# COVID-19 infections and outcomes in patients with multiple myeloma in New York City: a cohort study from five academic centers

**DOI:** 10.1101/2020.06.09.20126516

**Authors:** Malin Hultcrantz, Joshua Richter, Cara Rosenbaum, Dhwani Patel, Eric Smith, Neha Korde, Sydney Lu, Sham Mailankody, Urvi Shah, Alexander Lesokhin, Hani Hassoun, Carlyn Tan, Francesco Maura, Andriy Derkacs, Benjamin Diamond, Adriana Rossi, Roger N. Pearse, Deepu Madduri, Ajai Chari, David Kaminetsky, Marc Braunstein, Christian Gordillo, Faith Davies, Sundar Jagannath, Ruben Niesvizky, Suzanne Lentzsch, Gareth Morgan, Ola Landgren

## Abstract

**Importance:** New York City is a global epicenter for the SARS-CoV-2 outbreak with a significant number of individuals infected by the virus. Patients with multiple myeloma have a compromised immune system, due to both the disease and anti-myeloma therapies, and may therefore be particularly susceptible to coronavirus disease 2019 (COVID-19); however, there is limited information to guide clinical management.

**Objective:** To assess risk factors and outcomes of COVID-19 in patients with multiple myeloma.

**Design:** Case-series.

**Setting:** Five large academic centers in New York City.

**Participants:** Patients with multiple myeloma and related plasma cell disorders who were diagnosed with COVID-19 between March 10^th^, 2020 and April 30^th,^ 2020.

**Exposures:** Clinical features and risk factors were analyzed in relation to severity of COVID-19.

**Main Outcomes and Measures:** Descriptive statistics as well as logistic regression were used to estimate disease severity reflected in hospital admissions, intensive care unit (ICU) admission, need for mechanical ventilation, or death.

**Results:** Of 100 multiple myeloma patients (male 58%; median age 68, range 41-91) diagnosed with COVID-19, 74 (74%) were admitted; of these 13 (18%) patients were placed on mechanical ventilation, and 18 patients (24%) expired. None of the studied risk factors were significantly associated (P>0.05) with adverse outcomes (ICU-admission, mechanical ventilation, or death): hypertension (N=56) odds ratio (OR) 2.3 (95% confidence interval [CI] 0.9-5.9); diabetes (N=18) OR 1.1 (95% CI 0.3-3.2); age >65 years (N=63) OR 2.0 (95% CI 0.8-5.3); high dose melphalan with autologous stem cell transplant <12 months (N=7) OR 1.2 (95% CI 0.2-7.4), IgG<650 mg/dL (N=42) OR=1.2 (95% CI 0.4-3.1). In the entire series of 127 patients with plasma cell disorders, hypertension was significantly associated with the combined end-point (OR 3.4, 95% CI 1.5-8.1).

**Conclusions and Relevance:** Although multiple myeloma patients have a compromised immune system due to both the disease and therapy; in this largest disease specific cohort to date of patients with multiple myeloma and COVID-19, compared to the general population, we found risk factors for adverse outcome to be shared and mortality rates to be within the higher range of officially reported mortality rates.

## Introduction

The COVID-19 pandemic, caused by the SARS-CoV-2 virus, has become a global health crisis since it was first reported in Wuhan, China, in December 2019.^1^ COVID-19 has so far caused over 400,000 deaths globally and has spread to the majority of countries around the world.^2^ New York City is a global epicenter for the SARS-CoV-2 outbreak and a significant number of individuals have been infected by the virus, including both patients with underlying health conditions as well as healthy individuals.^3^ Clinical symptoms of COVID-19 include fever, cough, fatigue, diarrhea, headaches, and shortness of breath.^1^ They range from mild symptoms to severe disease characterized by pneumonia, hypoxia, respiratory failure, acute respiratory disease syndrome (ARDS), immune dysregulation, cytokine storm, thromboembolic events, and multiorgan failure.^1^ Reported risk factors for severe COVID-19 disease are male gender, advanced age, smoking, and certain comorbidities such as hypertension.^1,4^

Five studies of varying size have suggested that patients with cancer on active therapy or recent surgery had a higher risk of a more severe COVID-19 disease course.^5–9^ Additionally, recent immunotherapy treatment with checkpoint inhibitors was associated with a poorer outcome.^7^ Patients with multiple myeloma have an inherently compromised humoral and cellular immunity from the malignant plasma cell disorder itself and its associated hypogammaglobulinemia.^10^ The immunosuppression seen at presentation can be exacerbated by the standard combination anti-myeloma therapies currently in use.^11^ Among the conventional treatment options for multiple myeloma, the use of high-dose melphalan chemotherapy followed by autologous stem cell transplant is particularly associated with acute and sustained hypogammaglobulinemia and T-cell suppression.^12^ Here, we report on the largest experience to date from a cohort of multiple myeloma patients with COVID-19 from five large academic centers in New York City.

## Methods

Consecutive patients with multiple myeloma and related precursor diseases, hospitalized as well as outpatients, were included in this study. Participating centers are: Memorial Sloan Kettering Cancer Center (N=52), New York University Langone Health (N=30), Mount Sinai (N=23), Weill Cornell Medicine (N=13), and Columbia University Medical Center (N=9). The presence of SARS-CoV-2 was confirmed using real time polymerase chain reaction through nasopharynx swab. Patients with confirmed COVID-19 during the peak of the outbreak between March 10^th^ and April 30^th^, 2020 were included in this cohort study.

We obtained data on patient characteristics, comorbidities, laboratory findings, treatments, and outcomes. In addition to descriptive statistics, logistic regression was used to estimate risk factors associated with adverse outcomes. Taking this approach, we estimated odds ratios (ORs) with 95% confidence intervals (CIs). The primary composite end-point was admission to an intensive care unit (ICU), need for mechanical ventilation, or death. Hypogammaglobulinemia was defined as immunoglobulin G (IgG) levels <650 mg/dL (lower limit of normal), and severe hypogammaglobulinemia was defined as IgG levels <400 mg/dL. Separate analyses were performed for patients with multiple myeloma (N=100) and for all patients with plasma cell disorders (N=127).

## Results

We identified a total of 100 patients with multiple myeloma and COVID-19. Median age at the time of COVID-19 infection was 68 years (range 41-91 years). Fifty-eight (58%) of the patients were male and 24 were current or former smokers (**Table 1**). Concomitant cardiovascular or pulmonary comorbidities were seen in 74 patients of which hypertension was the most common (56%). Additionally, 27 patients with related plasma cell disorders; 20 with monoclonal gammopathy of undetermined significance (MGUS) and 3 with smoldering multiple myeloma (SMM), 3 patients with AL amyloidosis, and 1 patient with solitary plasmacytoma, were identified (**Table 1**).

**Table 1.** Patients’ characteristics.

|  | Number (%) |
| --- | --- |
| **All patients** | 127 (100) |
| **Men** | 68 (54) |
| **Women** | 59 (46) |
| **Median age at COVID-19 (years)** | 68 years |
| **Former/Current smoker** | 34 (27) |
| **Never smoker** | 92 (73) |
| **Multiple myeloma** | 100 (79) |
| **Newly diagnosed multiple myeloma** | 30 |
| **Stable multiple myeloma without relapse** | 35 |
| **Relapse/refractory multiple myeloma** | 35 |
| **Monoclonal gammopathy of undetermined significance** | 20 (16) |
| **Smoldering multiple myeloma** | 3 (2.4) |
| **AL amyloidosis** | 3 (2.4) |
| **Solitary plasmacytoma** | 1 (0.8) |

Three multiple myeloma patients had concomitant AL amyloidosis.

Of the 100 patients with multiple myeloma, 28 patients (28%) had newly diagnosed multiple myeloma; 26 were being treated with induction therapy, one had not yet started induction and one had chosen not to start therapy. There were 35 (35%) patients with stable disease (i.e. non-active disease); 23 of these were on lenalidomide maintenance, 5 were on other forms of maintenance including ixazomib/dexamethasone and daratumumab/dexamethasone and 7 patients with stable disease were monitored off therapy. Thirty-five (35%) patients had relapse/refractory multiple myeloma of which 31 were on various treatments for relapsed/refractory multiple myeloma; the majority daratumumab- or carfilzomib-based. The remaining 4 relapse/refractory patients were not on active anti-myeloma therapy due to advance disease stage (hospice) or other more pressing comorbidities. Information on treatment status was missing for two patients, these were both classified as newly diagnosed multiple myeloma.(**Table 2**) Overall, a total of 39 patients had undergone high-dose melphalan followed by autologous stem cell transplant; 7 patients within the 12 months prior to contracting COVID-19. Two patients had a prior allogeneic stem cell transplant several years before the COVID-19 diagnosis. (**Table 2**) Forty-two patients (42%) had hypo-gammaglobulinemia (IgG <650 mg/dL) and 18 patients (18%) had severe hypogammaglobulinemia (IgG <400 mg/dL).

**Table 2.** Treatment regimens in patients with multiple myeloma at the time of COVID-19 diagnosis.

|  | Number |
| --- | --- |
| **All patients** | 100 |
| **Patients with ongoing treatment** | 86 |
| **Bortezomib-including regimen** | 20 |
| **Carfilzomib-including regimen** | 15 |
| **Daratumumab-including regimen** | 24 |
| **Ixazomib-including regimen** | 6 |
| **Lenalidomide maintenance** | 22 |
| **Other treatments\*** | 10 |
| **Prior MEL/ASCT** | 39 |
| **Prior allogeneic transplant** | 2 |
| **Not on treatment** | 12 |
| **Missing information regarding treatment status** | 2 |

MEL/ASC=high dose melphalan and autologous stem cell transplantation
\*Other treatments included DCEP (dexamethasone, cyclophosphamide, etoposide, cisplatin), low dose melphalan, panabinostat, iberomid, chlorithromycin, venetoclax, selinexor, and AMG-701.

The laboratory findings in this multiple myeloma cohort revealed lymphopenia and elevated C-reactive protein, ferritin, D-dimer, and IL-6 levels. (**Table 3**) Patients who met the adverse combined endpoint (i.e. ICU-admission, mechanical ventilation, or death) had overall higher levels of inflammatory markers reflecting a more severe infection and cytokine activation.

**Table 3.**
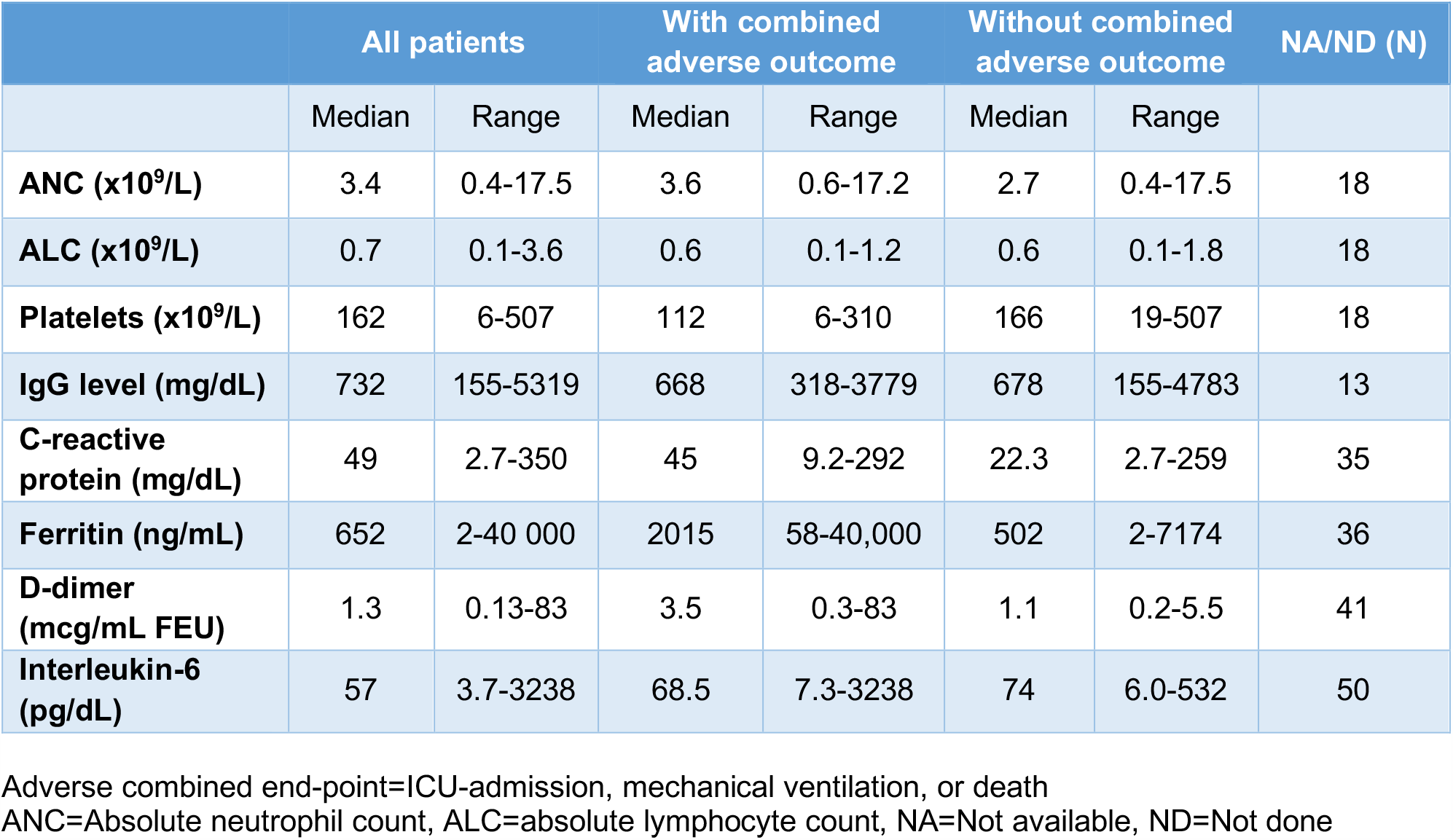
Laboratory findings in patients with multiple myeloma and COVID-19.

|  | All patients |  | With combined adverse outcome |  | Without combined adverse outcome |  | NA/ND (N) |
| --- | --- | --- | --- | --- | --- | --- | --- |
|  | Median | Range | Median | Range | Median | Range |  |
| **ANC (x10<sup>9</sup>/L)** | 3.4 | 0.4-17.5 | 3.6 | 0.6-17.2 | 2.7 | 0.4-17.5 | 18 |
| **ALC (x10<sup>9</sup>/L)** | 0.7 | 0.1-3.6 | 0.6 | 0.1-1.2 | 0.6 | 0.1-1.8 | 18 |
| **Platelets (x10<sup>9</sup>/L)** | 162 | 6-507 | 112 | 6-310 | 166 | 19-507 | 18 |
| **IgG level (mg/dL)** | 732 | 155-5319 | 668 | 318-3779 | 678 | 155-4783 | 13 |
| **C-reactive protein (mg/dL)** | 49 | 2.7-350 | 45 | 9.2-292 | 22.3 | 2.7-259 | 35 |
| **Ferritin (ng/mL)** | 652 | 2-40 000 | 2015 | 58-40,000 | 502 | 2-7174 | 36 |
| **D-dimer (mcg/mL FEU)** | 1.3 | 0.13-83 | 3.5 | 0.3-83 | 1.1 | 0.2-5.5 | 41 |
| **Interleukin-6 (pg/dL)** | 57 | 3.7-3238 | 68.5 | 7.3-3238 | 74 | 6.0-532 | 50 |

Adverse combined end-point=ICU-admission, mechanical ventilation, or death
ANC=Absolute neutrophil count, ALC=absolute lymphocyte count, NA=Not available, ND=Not done

Of the 100 patients with multiple myeloma and a SARS-CoV-2 positive RNA PCR test on this study, 74 (74%) were admitted due to COVID-19; 16 were admitted to the ICU and 13 of these patients were placed on mechanical ventilation. Regarding treatments used for COVID-19, 52 patients were treated with hydroxychloroquine and 52 with azithromycin, 42 had the combination of the two and 35 received neither hydroxychloroquine nor azithromycin. (**Table 4)** Nine patients received treatment with IL-6 inhibitors, tocilixumab or sarilumab, and one patient was treated with the IL-1 inhibitor anakinra and the TNF-alpha inhibitor infliximab. Other treatments included high dose steroids, broad-spectrum antibiotics, and investigational anti-viral therapies such as lopinavir-ritonavir and remdesivir. Two patients were treated with convalescent plasma. Eighteen of the multiple myeloma patients, thus 24% of those admitted, expired during the follow-up time.

**Table 4.** Treatment administered for COVID-19 in patients with multiple myeloma.

| Treatment for COVID19 | Number |
| --- | --- |
| Total number of patients | 100 |
| Patient receiving therapy for COVID-19 | 65 |
| Hydroxychloroquine | 52 |
| Azithromycin | 52 |
| Combination hydroxychloroquine and azithromycin | 40 |
| IL-6 blockade | 9 |
| Lopinavir-ritonavir | 4 |
| Remdisivir | 1 |

Twenty-seven patients (27%) of the 100 multiple myeloma patients met the combined adverse end-point (ICU-admission, mechanical ventilation, or death) and the risk was highest for patients with hypertension. None of the studied risk factors were significantly associated (P>0.05) with adverse outcomes, including hypertension, OR 2.3 (95% CI 0.9-5.9); diabetes, OR 1.1 (95% CI 0.3-3.2); age >65 years, OR 2.0 (95% CI 0.8-5.3), or male gender, OR 0.9 (95% CI 0.4-2.1). Based on small numbers (N=7), patients who received high-dose melphalan chemotherapy followed with autologous stem cell transplant within 12 months prior to COVID-19 diagnosis were not significantly associated with the adverse combined endpoint, OR 1.2 (95% CI 0.2-7.4). Also, we did not observe any statistical association between the adverse combined endpoint and hypogammaglobulinemia (IgG <650 mg/dL); OR 0.9 (95% CI 0.3-2.5) or severe hypogammaglobulinemia (IgG <400 mg/dL); and 1.2 (95% CI 0.4-4.2), respectively. (**Table 5**)

**Table 5.** Odds ratios (ORs) of the combined adverse end-point (intensive care unit admission, mechanical ventilation, or death).

|  | Multiple myeloma patients (N=100)<br>OR (95% CI) | Multiple myeloma and related plasma cell disorders (N=127)<br>OR (95% CI) |
| --- | --- | --- |
| **Any cardiovascular disease** | 1.3 (0.5-3.8) | 1.6 (0.6-4.2) |
| **Hypertension** | 2.3 (0.9-5.9) | 3.5 (1.5-8.1) |
| **Diabetes** | 1.1 (0.3-3.3) | 0.8 (0.3-2.4) |
| **COPD/asthma** | 2.8 (0.2-45.9) | 4.3 (0.98-19.2) |
| **Male gender** | 0.9 (0.4-2.1) | 0.7 (0.3-1.5) |
| **Age >65 years** | 2.0 (0.8-5.3) | 1.8 (0.8-4.1) |
| **MEL/ASCT <12m vs >12m prior to COVID-19** | 1.2 (0.2-7.4) | NA |
| **IgG <650 mg/dL** | 1.2 (0.4-3.1) | NA |
| **IgG <400 mg/dL** | 1.2 (0.4-4.2) | NA |

COPD=chronic obstructive pulmonary disorder
MEL/ASCT=high dose melphalan and autologous stem cell transplantation
IgG=Immunoglobulin G level

In the entire case series of 127 patients with plasma cell disorders, the OR of severe outcome (ICU-admission, mechanical ventilation, or death) was significantly elevated for those with hypertension (OR 3.4, 95% 1.5-8.1) while diabetes, male gender, age >65 years, or chronic obstructive pulmonary disease/asthma were not significantly associated with the adverse combined end-point.

## Discussion

COVID-19 is a disease caused by the SARS-CoV-2 virus where, in the general population, the most severe outcomes are observed among elderly patients and patients with cardiovascular comorbidities.^3,13^ There is limited data on outcomes in patients with cancers particularly patients with hematological malignancies. Here, we present the first large case series of COVID-19 in patients with multiple myeloma, a plasma cell malignancy associated with a compromised immune system, due to both disease biology and anti-myeloma therapies. Consecutive patients with multiple myeloma and related precursor diseases with confirmed presence of SARS-CoV-2 between March 1^st^ and April 30^th^, 2020 from five large academic centers in New York City were included in this study. In the general population, the probability of dying from COVID-19 has been reported to be between 0.5-3% including all COVID-19 positive patients, and between 6-30% for hospitalized patients due to the virus.^1,3,13-16^ Here, we show that among 74 multiple myeloma patients admitted due to COVID-19, the mortality rate was 24% which thus is in the higher range of what has been reported in the general population.

We were motivated to better understand risk factors associated with severe outcomes from COVID-19 positive multiple myeloma patients. Specifically, we studied host characteristics as well as available variables related to the disease and treatment. Similar to reports from the general population, we found more men than women diagnosed with COVID-19 in this series, and there was a tendency towards a higher OR for adverse outcome (ICU-admission, mechanical ventilation, or death) in patients with hypertension. In the analysis of the entire series of COVID-19 positive patients with multiple myeloma and related plasma cell disorders (N=127), similar to the general population, cardiovascular risk factors were significantly associated with adverse outcome.^3^ Independent of COVID-19, typically, patients with multiple myeloma are at an increased risk of infections due to both disease and treatment associated immunosuppression.^17^ Examples of these associations are the immunomodulatory and conventional chemotherapy drugs which cause myelosuppression, the targeted monoclonal antibodies such as anti-CD-38 associated with suppression of the humoral immune system.^18–22^ In the current series, we did not identify an association between hypogammaglobulinemia and adverse outcome; however, due to the lack of detailed biomarker data, we were unable to rule out associations involving other immunological mechanisms (such as T-cell suppression) that are important in the immune response towards viral and other infections. In multiple myeloma patients undergoing high-dose melphalan chemotherapy followed by autologous stem cell transplant, it is well-known that there are severe acute as well as sustained long-term immunosuppression including broad aspects of the immune system.^23,24^ In this study, 39 of the 100 multiple myeloma patients included had been treated with autologous stem cell transplant, 7 within the 12 months prior to contracting SARS-CoV-2. Based on small numbers (N=7), the OR for the adverse combined end-point was not significantly elevated in patients who had high-dose melphalan chemotherapy followed by autologous stem cell transplant within 12 months prior to contracting COVID-19. Despite not seeing an increased mortality associated with high-dose melphalan chemotherapy followed by autologous stem cell transplant, we note the concerns of the American Society of Hematology as well as the International Myeloma Society whose recommendations state that for transplant-eligible patients, high-dose melphalan chemotherapy followed with autologous stem cell transplant should be postponed, if possible, until the pandemic abates.^25,26^ Given that it is unknown whether there could be additional seasonal outbreaks in upcoming winter of 2020 as well as following years, it is good clinical practice to have this discussion with every patient. When a safe and efficacious SARS-CoV-2 vaccine becomes available, it should be recommended to multiple myeloma patients and should be added to re-immunization programs for melphalan-induced inactivation of prior vaccines.

To minimize the risk for exposure, management of multiple myeloma including the use of infusions, injections and oral drugs has been adjusted to favor fewer visits, fewer injections/infusions and a preference for oral drugs.^25–31^ All five academic centers limited the use of high-dose melphalan and autologous stem cell transplant and only a few high risk patients have been transplanted during the outbreak. Additionally, in our experience, patients with multiple myeloma have been particularly adherent to the general recommendations for social distancing.^32,33^ This may have contributed to the relatively low number of patients with multiple myeloma and COVID-19 in this cohort given the population of New York City (8.4 million), the overall number of confirmed cases (>200,000), and the large catchment area covered by the five included hospital centers.

In prior reports from the general population, the immune response in patients with severe COVID-19 show a unique pattern with high IL-6, low HLA-DR expression, and dysregulation of lymphocytes characterized by CD4 lymphopenia and subsequently B-cell lymphopenia.^34^ In the current series of multiple myeloma patients, we found high levels of IL-6, elevated ferritin, and low absolute lymphocyte levels. Furthermore, previous studies have reported that COVID-19 infection can lead to a hypercoagulable state and patients with severe COVID-19 have an increased risk of venous thromboembolism and stroke.^4,35^ In this series, D-dimer levels were found to be elevated and one patient had a cerebrovascular event leading to significant neurological sequelae. The biological underpinnings of these observations remain to be further explained in functional studies.

No treatment has so far shown an unequivocal beneficial effect for treatment of COVID-19. Hydroxychloroquine in combination with azithromycin was initially suggested to be an effective treatment combination; however, validation trials have been unable to confirm beneficial effects. Importantly, the World Health Organization recently expressed concern over cardiac arrhythmias and other potentially serious effects with the hydroxychloroquine-azithromycin combination and several of the initial publications have now been retracted.^36–39^ A few patients in our series were treated with lopinavir-ritonavir; however, clinical trials have not shown a clinical benefit of these anti-retroviral drugs.^40^ Recent reports on remdesivir as well as treatment with convalescent plasma therapy have showed promising results and there are case reports on successful treatment of COVID-19 with IL-6 blockade.^41–46^ Additionally, there are indications that the Bruton tyrosine kinase inhibitor acalabrutinib can reduce the excessive inflammatory response in patients with severe COVID-19.^47^

Limitations of this study includes it being a case-series from tertiary cancer centers with a selected patient population in which patients who were treated at local hospitals in the outpatient setting were less likely to be included. There was missing data for certain laboratory results for some of the patients, primarily those who were treated as outpatients where not all tested for C-reactive protein, ferritin, D-dimer, or IL-6 levels.

In summary, we present data on a large case-series of COVID-19 positive patients with multiple myeloma and related precursor diseases showing case fatality rates in the higher range of reports from the general population. We comprehensively investigated the role of other comorbidities and found that the strongest risk factors for severe outcome were similar to those in the general population. The molecular and immunological mechanisms responsible for this finding remain unclear; however, given that patients with multiple myeloma are at an increased risk of various other infections due to both disease and treatment associated immunosuppression ^17^, current recommendation by American Society of Hematology and the International Myeloma Society state that high-dose melphalan chemotherapy followed with autologous stem cell transplant should be postponed, if possible, until the pandemic levels off.^25,26^

Ongoing larger studies with a wide range of hematological malignancies will provide additional information on geographical variations of risk factors in these patients. Going forward, until there is a vaccine or effective treatment for COVID-19, clinical management and treatment of patients with multiple myeloma have to be carefully considered and adjusted to reduce the risk of exposure and minimize immune-suppression, while still aiming to achieve deep remissions in the era of COVID-19.

## Data Availability

Please contact the corresponding authors for questions regarding clinical data.

## Author Contributions

Dr. Hultcrantz had full access to all of the data in the study and takes responsibility for the integrity of the data and the accuracy of the data analysis.

Drs. Jagannath, Niesvizky, Lentzsch, Morgan and Landgren equally contributed to this work.

*Concept and design:* Hultcrantz, Derkach, Landgren

*Acquisition, analysis, or interpretation of data:* All authors

*Drafting of the manuscript:* Hultcrantz, Landgren

*Statistical analysis:* Hultcrantz, Derkach.

*Critical revision of the manuscript for important intellectual content:* All authors

## References

1. Guan WJ, Ni ZY, Hu Y, et al. Clinical Characteristics of Coronavirus Disease 2019 in China. N Engl J Med. 2020;382(18):1708–1720.

2. Center for Systems Science and Engineering (CSSE) at Johns Hopkins University B, MD, USA,. COVID-19 Dashboard. https://coronavirus.jhu.edu/map.html. Published 2020. Accessed 6/7/2020.

3. Richardson S, Hirsch JS, Narasimhan M, et al. Presenting Characteristics, Comorbidities, and Outcomes Among 5700 Patients Hospitalized With COVID-19 in the New York City Area. JAMA. 2020.

4. Oxley TJ, Mocco J, Majidi S, et al. Large-Vessel Stroke as a Presenting Feature of Covid-19 in the Young. N Engl J Med. 2020.

5. Liang W, Guan W, Chen R, et al. Cancer patients in SARS-CoV-2 infection: a nationwide analysis in China. Lancet Oncol. 2020;21(3):335–337.

6. He W, Chen L, Chen L, et al. COVID-19 in persons with haematological cancers. Leukemia. 2020.

7. Robilotti EV, Babady NE, Mead PA, et al. Determinants of Severity in Cancer Patients with COVID-19 Illness. medRxiv. 2020:2020.2005.2004.20086322.

8. Zhang L, Zhu F, Xie L, et al. Clinical characteristics of COVID-19-infected cancer patients: a retrospective case study in three hospitals within Wuhan, China. Ann Oncol. 2020.

9. Dai M, Liu D, Liu M, et al. Patients with Cancer Appear More Vulnerable to SARS-COV-2: A Multicenter Study during the COVID-19 Outbreak. Cancer Discov. 2020.

10. Heaney JLJ, Campbell JP, Iqbal G, et al. Characterisation of immunoparesis in newly diagnosed myeloma and its impact on progression-free and overall survival in both old and recent myeloma trials. Leukemia. 2018;32(8):1727–1738.

11. Kumar SK, Callander NS, Hillengass J, et al. NCCN Guidelines Insights: Multiple Myeloma, Version 1.2020. Journal of the National Comprehensive Cancer Network : JNCCN. 2019;17(10):1154–1165.

12. Steingrimsdottir H, Gruber A, Bjorkholm M, Svensson A, Hansson M. Immune reconstitution after autologous hematopoietic stem cell transplantation in relation to underlying disease, type of high-dose therapy and infectious complications. Haematologica. 2000;85(8):832–838.

13. Docherty AB, Harrison EM, Green CA, et al. Features of 16,749 hospitalised UK patients with COVID-19 using the ISARIC WHO Clinical Characterisation Protocol. medRxiv. 2020:2020.2004.2023.20076042.

14. Mehra MR, Desai SS, Kuy S, Henry TD, Patel AN. Cardiovascular Disease, Drug Therapy, and Mortality in Covid-19. N Engl J Med. 2020.

15. Goyal P, Choi JJ, Pinheiro LC, et al. Clinical Characteristics of Covid-19 in New York City. N Engl J Med. 2020.

16. Grasselli G, Zangrillo A, Zanella A, et al. Baseline Characteristics and Outcomes of 1591 Patients Infected With SARS-CoV-2 Admitted to ICUs of the Lombardy Region, Italy. JAMA. 2020.

17. Blimark C, Holmberg E, Mellqvist UH, et al. Multiple myeloma and infections: a population-based study on 9253 multiple myeloma patients. Haematologica. 2015;100(1):107–113.

18. Palumbo A, Cavallo F, Gay F, et al. Autologous transplantation and maintenance therapy in multiple myeloma. N Engl J Med. 2014;371(10):895–905.

19. Cavo M, Gay F, Beksac M, et al. Autologous haematopoietic stem-cell transplantation versus bortezomib-melphalan-prednisone, with or without bortezomib-lenalidomide-dexamethasone consolidation therapy, and lenalidomide maintenance for newly diagnosed multiple myeloma (EMN02/HO95): a multicentre, randomised, open-label, phase 3 study. Lancet Haematol. 2020.

20. Nahi H, Chrobok M, Gran C, et al. Infectious complications and NK cell depletion following daratumumab treatment of Multiple Myeloma. PLoS One. 2019;14(2):e0211927.

21. Mateos MV, Dimopoulos MA, Cavo M, et al. Daratumumab plus Bortezomib, Melphalan, and Prednisone for Untreated Myeloma. N Engl J Med. 2018;378(6):518–528.

22. D’Agostino M, Boccadoro M, Smith EL. Novel Immunotherapies for Multiple Myeloma. Curr Hematol Malig Rep. 2017;12(4):344–357.

23. Fermand JP, Katsahian S, Divine M, et al. High-dose therapy and autologous blood stem-cell transplantation compared with conventional treatment in myeloma patients aged 55 to 65 years: long-term results of a randomized control trial from the Group Myelome-Autogreffe. J Clin Oncol. 2005;23(36):9227–9233.

24. Attal M, Lauwers-Cances V, Hulin C, et al. Lenalidomide, Bortezomib, and Dexamethasone with Transplantation for Myeloma. N Engl J Med. 2017;376(14):1311–1320.

25. American Society of Hematology. COVID-19 and Multiple Myeloma. https://www.hematology.org/covid-19/covid-19-and-multiple-myeloma. Published 2020. Accessed 5/8/2020.

26. International Myeloma Society. International Myeloma Society Recommendations for the Management of Myeloma Patients During the COVID-19 Pandemic. 2020.https://cms.cws.net/content/beta.myelomasociety.org/files/IMS%20recommendations%20for%20Physicians%20Final.pdf Accessed 5/8/2020.

27. Al Saleh AS, Sher T, Gertz MA. Multiple Myeloma in the Time of COVID-19. Acta Haematol. 2020:1–7.

28. Malard F, Mohty M. Management of patients with multiple myeloma during the COVID-19 pandemic. Lancet Haematol. 2020.

29. National Comprehenstive Cancer Network. NCCN Coronavirus Disease 2019 (COVID-19) Resources for the Cancer Care Community 2020 at https://www.nccn.org/covid-19/ Accessed 5/8/2020.

30. American Society of Clinical Oncology. ASCO Coronavirus Resources. 2020 at https://www.asco.org/asco-coronavirus-information Accessed 5/8/2020

31. European Society for Medical Oncology. Cancer Patient Management During the COVID-19 Pandemic. 2020 at https://www.esmo.org/guidelines/cancer-patient-management-during-the-covid-19-pandemic Accessed 5/8/2020

32. Gudbjartsson DF, Helgason A, Jonsson H, et al. Spread of SARS-CoV-2 in the Icelandic Population. N Engl J Med. 2020.

33. Pan A, Liu L, Wang C, et al. Association of Public Health Interventions With the Epidemiology of the COVID-19 Outbreak in Wuhan, China. JAMA. 2020.

34. Giamarellos-Bourboulis EJ, Netea MG, Rovina N, et al. Complex Immune Dysregulation in COVID-19 Patients with Severe Respiratory Failure. Cell Host Microbe. 2020.

35. Klok FA, Kruip M, van der Meer NJM, et al. Confirmation of the high cumulative incidence of thrombotic complications in critically ill ICU patients with COVID-19: An updated analysis. Thromb Res. 2020.

36. Chang D, Saleh M, Gabriels J, et al. Inpatient Use of Ambulatory Telemetry Monitors for COVID-19 Patients Treated with Hydroxychloroquine and/or Azithromycin. J Am Coll Cardiol. 2020.

37. Gautret P, Lagier JC, Parola P, et al. Clinical and microbiological effect of a combination of hydroxychloroquine and azithromycin in 80 COVID-19 patients with at least a six-day follow up: A pilot observational study. Travel Med Infect Dis. 2020:101663.

38. Geleris J, Sun Y, Platt J, et al. Observational Study of Hydroxychloroquine in Hospitalized Patients with Covid-19. N Engl J Med. 2020.

39. Mehra MR, Desai SS, Ruschitzka F, Patel AN. Hydroxychloroquine or chloroquine with or without a macrolide for treatment of COVID-19: a multinational registry analysis. Lancet. 2020.

40. Cao B, Wang Y, Wen D, et al. A Trial of Lopinavir-Ritonavir in Adults Hospitalized with Severe Covid-19. N Engl J Med. 2020.

41. Zhang X, Song K, Tong F, et al. First case of COVID-19 in a patient with multiple myeloma successfully treated with tocilizumab. Blood Advances. 2020;4(7):1307–1310.

42. Shen C, Wang Z, Zhao F, et al. Treatment of 5 Critically Ill Patients With COVID-19 With Convalescent Plasma. JAMA. 2020.

43. Grein J, Ohmagari N, Shin D, et al. Compassionate Use of Remdesivir for Patients with Severe Covid-19. N Engl J Med. 2020.

44. Chaidos A, Katsarou A, Mustafa C, Milojkovic D, Karadimitris A. IL-6 blockade treatment for severe COVID-19 in two patients with multiple myeloma. Br J Haematol. 2020.

45. Beigel JH, Tomashek KM, Dodd LE, et al. Remdesivir for the Treatment of Covid-19 - Preliminary Report. N Engl J Med. 2020.

46. Li L, Zhang W, Hu Y, et al. Effect of Convalescent Plasma Therapy on Time to Clinical Improvement in Patients With Severe and Life-threatening COVID-19: A Randomized Clinical Trial. JAMA. 2020.

47. Roschewski M, Lionakis MS, Sharman JP, et al. Inhibition of Bruton tyrosine kinase in patients with severe COVID-19. Sci Immunol. 2020;5(48).

